# Plasma lipidomics identifies a signature of NAFLD in children that couples with cardiometabolic outcomes in adults

**DOI:** 10.1101/2020.04.18.20070417

**Authors:** Jake P. Mann, Benjamin Jenkins, Samuel Furse, Stuart G. Snowden, Anna Alisi, Laura G. Draijer, EU-PNAFLD investigators, Kylie Karnebeek, Deirdre A. Kelly, Bart G. Koot, Antonella Mosca, Camilla Salvestrini, Indra van Mourik, Anita Vreugdenhil, Matthias Zilbauer, Albert Koulman

## Abstract

Non-alcoholic fatty liver disease (NAFLD) is an increasingly common condition in children and adults characterized by insulin resistance and altered lipid metabolism. Affected patients are at increased risk of cardiovascular disease (CVD) and children with NAFLD are likely to be at risk of premature cardiac events. Evaluation of the plasma lipid profile of children with NAFLD offers the opportunity to investigate these perturbations and understand how closely they mimic the changes seen in adults with cardiometabolic disease. We hypothesized that change in the concentration of lipid species in pediatric NAFLD would mimic the alterations known to be associated with CVD in adults (and be largely reflective of insulin resistance). Here, we performed untargeted liquid chromatography mass spectrometry (LC-MS) plasma lipidomics on 287 children: 19 lean controls, 146 from an obese cohort, and 122 NAFLD cases who had undergone liver biopsy. Associations between lipid species and liver histology were assessed using regression adjusted for age and sex. Results were then replicated using data from 9,500 adults with metabolic phenotyping. Phosphatidylcholine (PC) and triglyceride (TG) desaturation and chain length were inversely associated with histological severity of paediatric NAFLD. Nine lipids species (lyso-PC, PC, and TG classes) were also associated with hepatic steatosis and insulin resistance in an independent cohort of adults. Five of the 9 lipids replicated in the adults cohort (including PC(36:4)) have been directly linked to death and cardiometabolic disease in adults, as well as indirectly via genetic variants that influence the concentration of these species. Together, these findings suggest that lipid metabolism is altered in paediatric NAFLD in a similar way as in cardiometabolic disease in adults and it is therefore critical to alleviate insulin resistance in these children to mitigate their long-term morbidity.

## Introduction

Non-alcoholic fatty liver disease (NAFLD) is a common, chronic disorder that is closely linked to obesity and insulin resistance[1]. Most of the morbidity and mortality in patients with NAFLD occurs due to complications of cardiovascular disease though a small, but significant, proportion develop cirrhosis[2]. Individuals with higher stages of liver fibrosis[3] or more active inflammation (non-alcoholic steatohepatitis (NASH)[4] are more likely to progress to end-stage liver disease and may be at higher risk of cardiovascular disease CVD[5].

The current increase in childhood obesity is resulting to more NAFLD in children, which is also associated with the metabolic syndrome[6] and, whilst the long-term outcomes have not yet been formally established with the same degree of confidence as in adults[7,8], they are believed to be similar, including cardiovascular disease (CVD). However, paediatric NAFLD has several unique features, in particular peri-portal inflammation is a more prominent feature, particularly in younger children, boys, and in those of Hispanic ethnicity[9,10]. Therefore, it is not entirely clear to what extent paediatric and adult NAFLD differ.

Lipidomics is a technique that aims to comprehensively measure the concentration (or abundance) of lipid species that has been used by several groups to gain insight into altered lipid metabolism in NAFLD. Liver samples[11–13], venous-[14,15] and portal-blood[16] have been studied in humans, all showing specific lipid species to associate with histological severity of NAFLD in adults. This work has identified perturbation of pathways leading to an increased hepatic *de novo* lipogenesis, desaturase activity, and phospholipid metabolism. To date, lipidomic studies in children have focused on species that differentiate NAFLD patients from healthy or obese controls [17,18]. However, none of these studies have included histologically characterized cases therefore it is not clear whether the observed changes are purely reflective of underlying insulin resistance or specific to NAFLD. and its histological severity.

In this study we analyzed plasma lipidomics to investigate lipid metabolism in children with NAFLD. Specifically, we aimed to: (i) identify lipid species that were associated with the histological severity of NAFLD; (ii) determine if similar changes were observed in separate cohort of obese children; (iii) identify overlap in a cohort of adults with NAFLD; (iv) and then to understand the potential long-term clinical significance of these lipids on cardiometabolic disease outcomes using data from adults. We hypothesized that the lipid signature of paediatric NAFLD would be largely reflective of insulin resistance and therefore would be associated with cardiometabolic disease in adults.

## Methods

### Participants

An overview of the study design is shown in Figure 1. Three groups of participants were included in this cross-sectional study: lean controls, a cohort of children who were overweight or obese (‘obesity cohort’), and cases with suspected advanced NAFLD who had undergone liver biopsy (‘biopsied NAFLD cases’). In addition, we used publicly available data from adults cohorts [19–21]. All participants (or their parents) gave written informed consent and were recruited between 2014-2019, for the below ethically-approved studies, which were confirmed with the Declaration of Helsinki principles.

**Figure 1.**
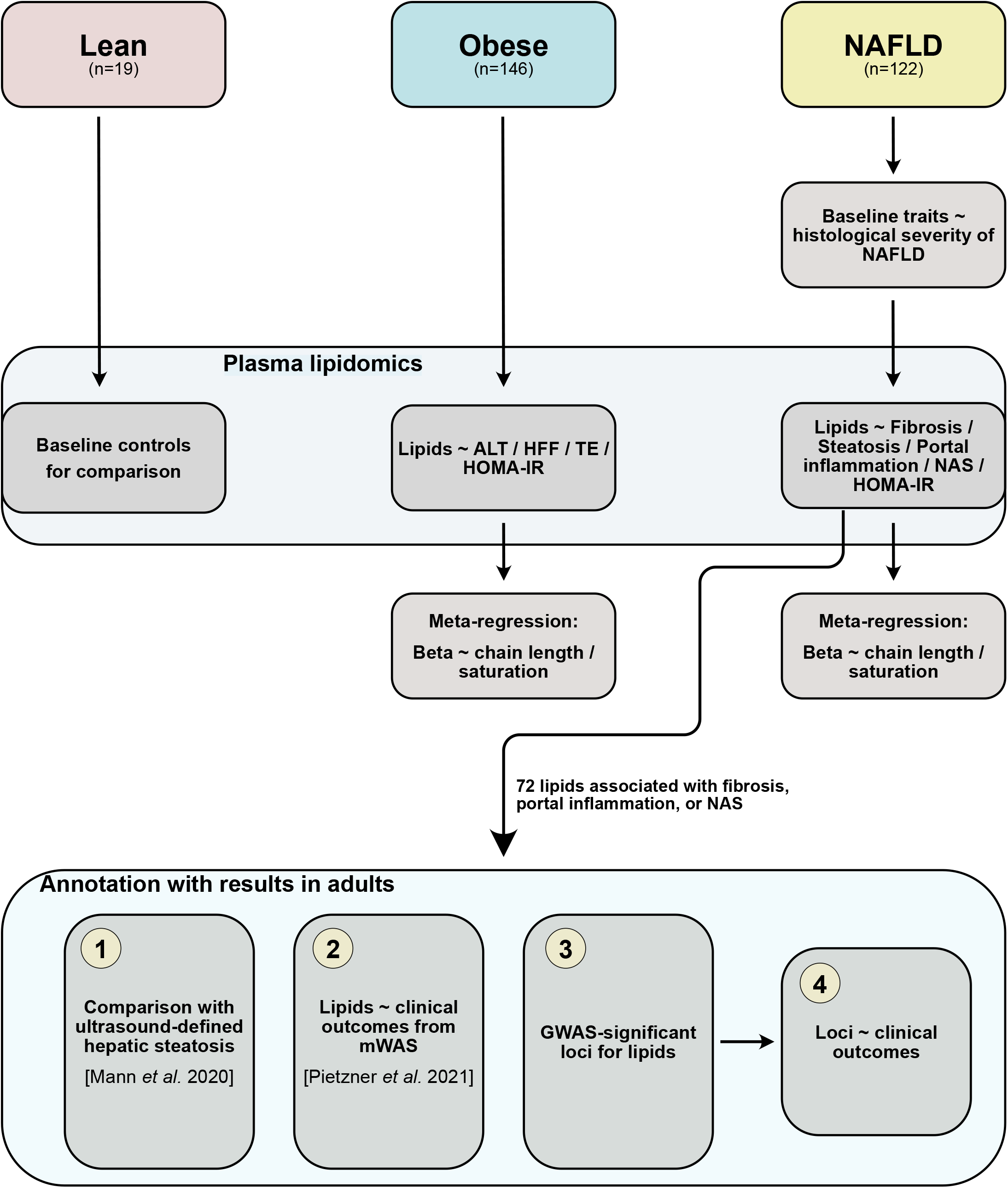
Overview of study design. Three groups of a participants were included: lean controls, an obesity cohort, and cases biopsied for NAFLD. The association between baseline characteristics and histological severity of NAFLD was tested using linear regression. Plasma lipidomics was run in all three groups and analyzed using linear regression for HOMA-IR and liver-related traits. Meta-regression was performed on results from the obesity cohort and biopsied NAFLD cases. The 72 lipids associated with severity of NAFLD were annotated with four sets of results from adults. ALT, alanine aminotransferase; HFF, hepatic fat fraction; HOMA-IR, homeostatic model of insulin resistance; mWAS, metabolite-wide association study; NAS, NAFLD Activity Score; TE, transient elastography.

Lean controls were recruited as part of the Translational Research in Intestinal Physiology and Pathology (TRIPP) Study at Cambridge University (UK), which was approved by East of England - Cambridge South Research Ethics Committee (REC 17/EE/0265). These children had been referred due to diarrhea, vomiting, or abdominal pain and underwent endoscopy to rule out gastrointestinal disease. They were found to have no evidence of pathology after thorough assessment and had complete resolution of any symptoms. There is a low likelihood of NAFLD is this control group who were lean (BMI z-score <1.04) and had normal liver biochemistry.

The obesity cohort was recruited from paediatric obesity clinics at Maastricht Children’s Hospital (under ethical approval METC 13-4-130) and Amsterdam University Medical Centers (under ethical approval MEC 2017_306 and MEC 07/141). Children were referred to these clinics from their primary care physicians due to being overweight or obese and were then subsequently investigated for co-morbidities (and secondary causes of obesity). As there was no clinical indication for liver biopsy, it was not possible to conclusively identify or exclude NAFLD in all children from the obesity cohort. However, a subset underwent magnetic resonance spectroscopy (MRS) which provides high sensitivity for identifying steatosis.

Biopsied NAFLD cases were recruited as part of the European Paediatric NAFLD Registry (EU-PNAFLD, Clintrials.gov NCT:04190849)[22], which was approved by the East Midlands - Nottingham 2 Research Ethics Committee (17/EM/0084). These children had been referred to specialist centres for paediatric hepatology (Birmingham Children’s Hospital (UK) and Bambino Gesù Children’s Hospital (Rome, Italy)) due to suspected advanced NAFLD and underwent liver biopsy for diagnosis of NAFLD and staging of disease.

As an exploratory analysis, we utilized data from the maximum number of available participants therefore no formal sample size calculation was performed.

### Inclusion and exclusion criteria

All participants were 5-18 years old at the time of inclusion.

Lean controls: BMI z-score <1.04, normal serum liver biochemistry, and no clinical evidence of liver disease. The obesity cohort all had a BMI z-score >1.64 (except for one participant who was overweight with BMI z-score 1.2) and no evidence of secondary causes of obesity (e.g. hypothyroidism). Biopsied NAFLD cases: NAFLD diagnosed by liver histology, plus exclusion of secondary causes using blood tests: normal range ceruloplasmin, ferritin, alpha-1-antitrypsin; normal immunoglobulins, negative antinuclear antibody, and negative anti-smooth muscle antibody; and negative screening for chronic viral hepatitis.

### Clinical and laboratory investigations

Lean controls had height and weight measured and fasting blood obtained but no further samples or investigations were performed for the purpose of this study.

The obesity cohort and biopsied NAFLD cases both underwent baseline characterization with clinical assessment and anthropometric measurements including waist/hip circumferences and blood pressure. Fasting blood was obtained and plasma was frozen at -80 degree Celsius for use in all analyses. The homeostatic model assessment of insulin resistance (HOMA-IR) was derived using fasting insulin (µU/L) x fasting glucose (nmol/L) / 22.5. A subset of the obesity cohort (n=95) underwent magnetic resonance spectroscopy (MRS) for quantification of hepatic fat and a separate subset (n=42) had transient elastography (TE) performed to non-invasively test for signs of NAFLD. Steatosis was defined as MRS >1.8%, which corresponds with >5% fat containing hepatocytes on liver histology[23].

### Liver biopsies

All NAFLD cases underwent liver biopsy whereas liver biopsy was not performed in control participants or the obesity cohort. Liver biopsy was performed in accordance with ESPGHAN guidelines[24]. Biopsies were obtained percutaneously and considered adequate if there was minimal fragmentation and at least 20 mm long. Liver biopsy was scored according to NASH CRN criteria[10,25], including assessment of portal inflammation as none (0), mild (1), or more than mild (2), and calculation of the NAFLD Activity Score (NAS). Samples from liver biopsy were used for histological assessment only; no tissue was available for lipidomic (or transcriptomic) analysis.

### Plasma lipidomics analysis

For lipid profiling, the plasma samples were analyzed by liquid chromatography with mass spectrometry detection (LC–MS) as described previously[26].

All solvents and additives were of HPLC grade or higher and purchased from Sigma Aldrich (Haverhill, Suffolk, UK) unless otherwise stated.

The protein□precipitation liquid extraction protocol has been described previously[26]. Briefly, 50 µL of plasma was transferred into a 2 mL screw cap Eppendorf plastic tube (Eppendorf, Stevenage, UK). Immediately, 650 µL of chloroform was added to each sample, followed by thorough mixing. Then, 100 µL of the LIPID-IS (5 µM in methanol), 100 µL of the CARNITINE-IS (5 µM in methanol) and 150 µL of methanol was added to each sample, followed by thorough mixing. Then, 400 µL of acetone was added to each sample. The samples were vortexed and centrifuged for 10 minutes at ∼20,000 g to pellet any insoluble material. The supernatant was pipetted into separate 2 mL screw cap amber-glass auto-sampler vials (Agilent Technologies, Cheadle, United Kingdom). The organic extracts were dried down to dryness using a Concentrator Plus system (Eppendorf, Stevenage, UK) run for 60 minutes at 60 degree Celsius. The samples were reconstituted in 100 µL of 2: 1: 1 (propan□2□ol, acetonitrile and water, respectively) then thoroughly vortex. The reconstituted sample was transferred into a 250 μL low-volume vial insert inside a 2 mL amber glass auto-sample vial ready for liquid chromatography with mass spectrometry detection (LC-MS) analysis.

Full chromatographic separation of intact lipids was achieved using Shimadzu HPLC System (Shimadzu UK Limited, Milton Keynes, United Kingdom) with the injection of 10 µL onto a Waters Acquity UPLC® CSH C18 column (Waters, Hertfordshire, United Kingdom); 1.7 µm, I.D. 2.1 mm × 50 mm, maintained at 55 degrees Celsius. Mobile phase A was 6:4, acetonitrile and water with 10 mM ammonium formate. Mobile phase B was 9:1, propan-2-ol and acetonitrile with 10 mM ammonium formate. The flow was maintained at 500 µL per minute through the following gradient: 0.00 minutes_40% mobile phase B; 0.40 minutes_43% mobile phase B; 0.45 minutes_50% mobile phase B; 2.40 minutes_54% mobile phase B; 2.45 minutes_70% mobile phase B; 7.00 minutes_99% mobile phase B; 8.00 minutes_99% mobile phase B; 8.3 minutes_40% mobile phase B; 10 minutes_40% mobile phase B. The sample injection needle was washed using 9:1, 2-propan-2-ol and acetonitrile. The mass spectrometer used was the Thermo Scientific Exactive Orbitrap with a heated electrospray ionization source (Thermo Fisher Scientific, Hemel Hempstead, UK). The mass spectrometer was calibrated immediately before sample analysis using positive and negative ionization calibration solution (recommended by Thermo Scientific). Additionally, the mass spectrometer scan rate was set at 4 Hz, giving a resolution of 25,000 (at 200 m/z) with a full-scan range of m/z 100 to 1,800 with continuous switching between positive and negative mode.

#### Data processing

The instrument responses of the analytes were normalized to the relevant internal standard response (producing area ratios), these area ratios corrected the intensity for any extraction and instrument variations. The area ratios were then blank corrected where intensities less than three times the blank samples were set to a ‘Not Found’ result (i.e., zero concentration). The accepted area ratios were then multiplied by the concentration of the internal standard to give the analyte semi-quantitative concentrations.

### Statistical analyses

Participants were initially separated into lean controls, obesity cohort, and biopsied NAFLD cases. Clinical and biochemical characteristics of obese controls and biopsied NAFLD cases were compared using two-way unpaired T-tests for continuous data and chi-squared test for categorical data. p-values were converted to q-values by adjusting for multiple testing using the Benjamini-Hochberg method. Due to the differences in clinical characteristics and selection of participants, further analyses were performed within either the obesity cohort or biopsied NAFLD cases separately.

In the NAFLD cohort, the association between standard clinical or biochemical measurements and liver histology was tested using regression analysis. Skewed continuous independent variables (e.g. age, HOMA-IR), were logarithmically transformed and standardized (to mean = 0, standard deviation (SD) = 1) before being used in linear models. Univariable linear regression was performed for NAS, fibrosis, and portal inflammation score (dependent variables) and each independent variable.

Following initial processing as described above, only lipid species detected in >70% of participants (including lean controls) were included. For included species, minimum value imputation was used for missing values. ‘Total’ values for each lipid class (e.g. ‘total lysophosphatidylcholine (LPC)’) were calculated as the sum of all species within a class for each individual. Absolute concentrations of lipids were logarithmically transformed and standardized (to mean = 0, SD = 1) to account for skewed distributions and allow linear regression analyses.

In the obesity cohort, linear regression was performed between lipid species and serum alanine aminotransferase (ALT), HOMA-IR, hepatic fat fraction (HFF) on MRS, and liver stiffness (kPa) measured by transient elastography. In the biopsied NAFLD cases, linear regression was performed between lipid species and NAS, steatosis grade, portal inflammation score, fibrosis stage, and HOMA-IR. All linear regressions were adjusted for age and sex. Due to high correlation between lipid species, the critical p-value for significance was defined by 0.05/√n, where n is the number of included lipids: n=229 therefore p<3.3×10^−3^ was determined as statistically significant.

We then performed meta-regression to examine for trends in lipid saturation or carbon chain length within classes of lipids. Beta regression coefficients from the above models were regressed against double bonds or carbons within lipid classes. p-values were converted to q-values by adjusting for multiple testing using the Benjamini-Hochberg method.

### Comparison with adult NAFLD

We explored whether significant, directionally consistent associations were observed between lipids identified in this study and those in a previously published cohort study of adults[19]. This study used both targeted and untargeted metabolomics in a population-based study of middle-aged adults who had undergone abdominal ultrasound for identification of hepatic steatosis, in addition to comprehensive metabolic analysis. Further details of the Fenland Cohort are described elsewhere[27]. We searched for all directionally consistent and significant (q<0.1) associations for the 72 lipids that reached statistical significance from our analyses in pediatric biopsied NAFLD cases.

### Annotation with disease outcomes and GWAS loci

The 72 significant lipids from this study were also annotated with results from a recent metabolite-wide association study of non-communicable diseases in adults[20]. This study used untargeted metabolomics in the EPIC-Norfolk cohort of adults with long-term prospective follow-up for clinical outcomes, as a component of the EPIC study[28]. We searched for all directionally consistent and significant (q<0.1) associations from Pietzner *et al*. for the 72 lipids identified from our analyses in biopsied NAFLD cases.

Next, we explored which genetic variants are known to influence circulating levels of the lipids identified in the current analysis. We used data from a lipidomics genome-wide association study (GWAS), which had also collated relevant results from previous metabolomics GWAS[21]. We searched for genome-wide significant variants (as defined by the original studies) for the 72 top lipids identified in our analysis. The nearest gene annotation was extracted from the original study and no functional annotation of variants was performed.

Finally, for variants at GWAS significance from the above data, we performed a phenome-wide association study for cardio-metabolic traits. We used PheWAS data from Tabassum *et al*. and searched PhenoscannerV2[29] to annotate variants.

All analyses were performed using R 4.0.2[30] and code used is available from: https://doi.org/10.5281/zenodo.5507129.

## Results

287 children were recruited to the study: 19 lean controls, 146 obese or overweight children, and 122 biopsied NAFLD cases (Table 1). We studied a cohort of obese children and a group of specifically selected children with NAFLD who had undergone biopsy in a tertiary liver center. These three groups were brought together to understand differences in circulating lipid profile in severe NAFLD and compared to other children with obesity more typical of those seen in primary care.

**Table 1.** Baseline characteristics of participants included in the study. Lean children (n=19) were those undergoing endoscopy who had no evidence of gastrointestinal pathology. Obesity cohort (n=146) comprised children who were overweight or obese referred and for clinical assessment. The NAFLD cases (n=122) were children with suspected severe paediatric NAFLD who underwent liver biopsy. q-values represent false-discovery rate correct p-values between obesity and NAFLD cohorts, using unpaired t-tests for continuous traits and chi-squared for sex. Data represents mean (standard deviation) for continuous traits and number (%) for categorical traits. ALP, alkaline phosphatase; ALT, alanine aminotransferase; AST, aspartate aminotransferase; BMI, body mass index; GGT, gamma-glutamyl transferase; HDL, high density lipoprotein; HOMA-IR, homeostatic model of insulin resistance; LDL, low density lipoprotein; MRS, magnetic resonance spectroscopy.

|  | **Overall** | **Lean**<br>**controls** | **Obese**<br>**cohort** | **NAFLD**<br>**cases** | **q-value** |
| --- | --- | --- | --- | --- | --- |
| n | 287 | 19 | 146 | 122 |  |
| Age (years) | 13.0 (2.8) | 11.0 (3.4) | 14.2 (2.4) | 12.0 (2.5) | 1.00E-10 |
| Female sex | 162 (52.4) | 10 (52.6) | 76 (52.1) | 76 (52.8) |  |
| BMI z-score | 2.48 (1.1) | -0.04 (1.2) | 3.16 (0.6) | 2.15 (0.6) | 5.00E-27 |
| HOMA-IR | 3.6 (2.2) |  | 3.8 (2.6) | 3.2 (1.7) | 0.74 |
| Hypertension, n/total<br>(%) | 74/233 (32) |  | 59/119 (50) | 15/114 (13) | 2.40E-9 |
| Triglycerides<br>(mmol/L) | 1.2 (0.7) |  | 1.0 (0.6) | 1.4 (0.7) | 1.60E-07 |
| Total cholesterol<br>(mmol/L) | 4.2 (0.8) |  | 4.0 (0.8) | 4.3 (0.9) | 0.35 |
| HDL cholesterol<br>(mmol/L) | 1.1 (0.3) |  | 1.1 (0.3) | 1.1 (0.3) | 1 |
| LDL cholesterol<br>(mmol/L) | 2.5 (0.6) |  | 2.5 (0.7) | 2.6 (0.5) | 0.07 |
| ALT (IU/L) | 55.3 (41.8) |  | 41.9 (37.4) | 69.3 (41.7) | 8.1E-13 |
| AST (IU/L) | 38.6 (20.7) |  | 30.0 (14.5) | 47.7 (22.3) | 2.3E-16 |
| GGT (IU/L) | 25.4 (12.8) |  | 24.3 (11.8) | 36.4 (17.3) | 9.0E-03 |
| ALP (IU/L) | 200.8 (122.3) |  | 184.3 (113.6) | 260.1<br>(139.7) | 0.66 |
| Hepatic fat fraction on<br>MRS (%) | 3.6 (4.7) |  | 3.6 (4.7) |  |  |

The obese cohort reflected referral from primary care physicians and therefore included a full spectrum of individuals from those with no evidence of systemic metabolic dysfunction, through to those with marked insulin resistance (Figure S1B). Of the subset from the obese cohort who undergone liver MRS, 47% (45/90) had steatosis (i.e. >1.8% on MRS, Figure S1F).

In the biopsied NAFLD cases, a range of histological severity was observed (Table S1). 55/114 (48%) had NAFLD Activity Score ≥5 and 86/115 (75%) had evidence of portal inflammation. 100/122 (82%) of children had fibrosis and, though 5 children had stage 3 fibrosis, none were cirrhotic.

Consistent with previous reports, typical liver-related biochemistry (i.e. ALT, AST) were poor predictors of histological severity of NAFLD and standard biochemical serum lipids (total triglycerides and total cholesterol) were also not associated NAS or portal inflammation. Only age (Figure 2A) and severity of insulin resistance (Figure 2C, as approximated using HOMA-IR) were associated with histological severity of NAFLD, for example HOMA-IR and NAS beta=1.2±.3, q=3.1×10^−3^, adj.r^2^=.12 (Table S2).

**Figure 2.**
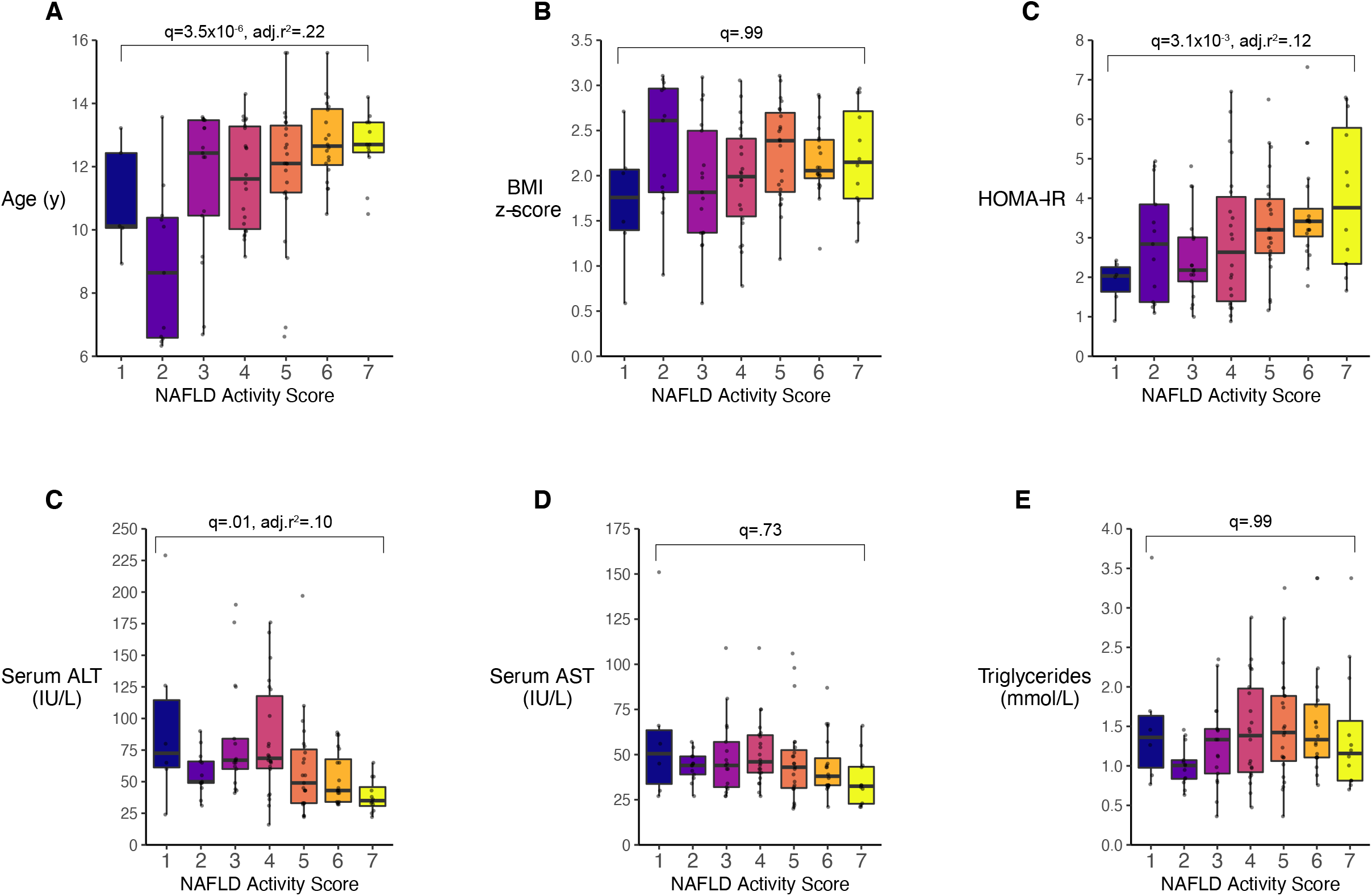
Associations between baseline characteristics and NAFLD Activity Score in children biopsied for NAFLD (n=122). Age (A), body mass index (BMI) z-score (B), and homeostatic model of insulin resistance (HOMA-IR, C) were positively associated with NAFLD Activity Score, whilst serum alanine aminotransferase (C) was negatively associated. Other aspartate aminotransferase (D) and total serum triglycerides (E) were not associated with NAFLD Activity Score. Associations were tested using linear regression. q-values were derived using the Benjamini-Hochberg method where significance is q<.05.

Therefore, we performed plasma lipidomics to investigate whether individual lipid species (or trends in lipid saturation / chain length) were associated with the severity of NAFLD. Following normalization, all cohorts overlapped on principal component analysis and there was no clear separation of obese vs NAFLD children by hierarchical clustering (Figure S3A).

Using linear regression, we tested the association between various metabolic and hepatic traits with each lipid species within either obese cohort or biopsied NAFLD cases (Table S3 & Figure S3C). Similar results were observed for HOMA-IR, NAS, and steatosis grade in the biopsied NAFLD cases, whilst HOMA-IR clustered with ALT and HFF% in the obesity cases. These groups of traits also demonstrated significant correlation with HOMA-IR in both groups (Figure S3E & F).

We observed consistent trends on meta-regression for multiple lipid classes across both cohorts (Table S4, Figure S3D), for example, serum ALT and hepatic fat fraction (in obese cohort), and NAS (in NAFLD cohort) were negatively associated with TG desaturation and chain length (Figure 3).

**Figure 3.**
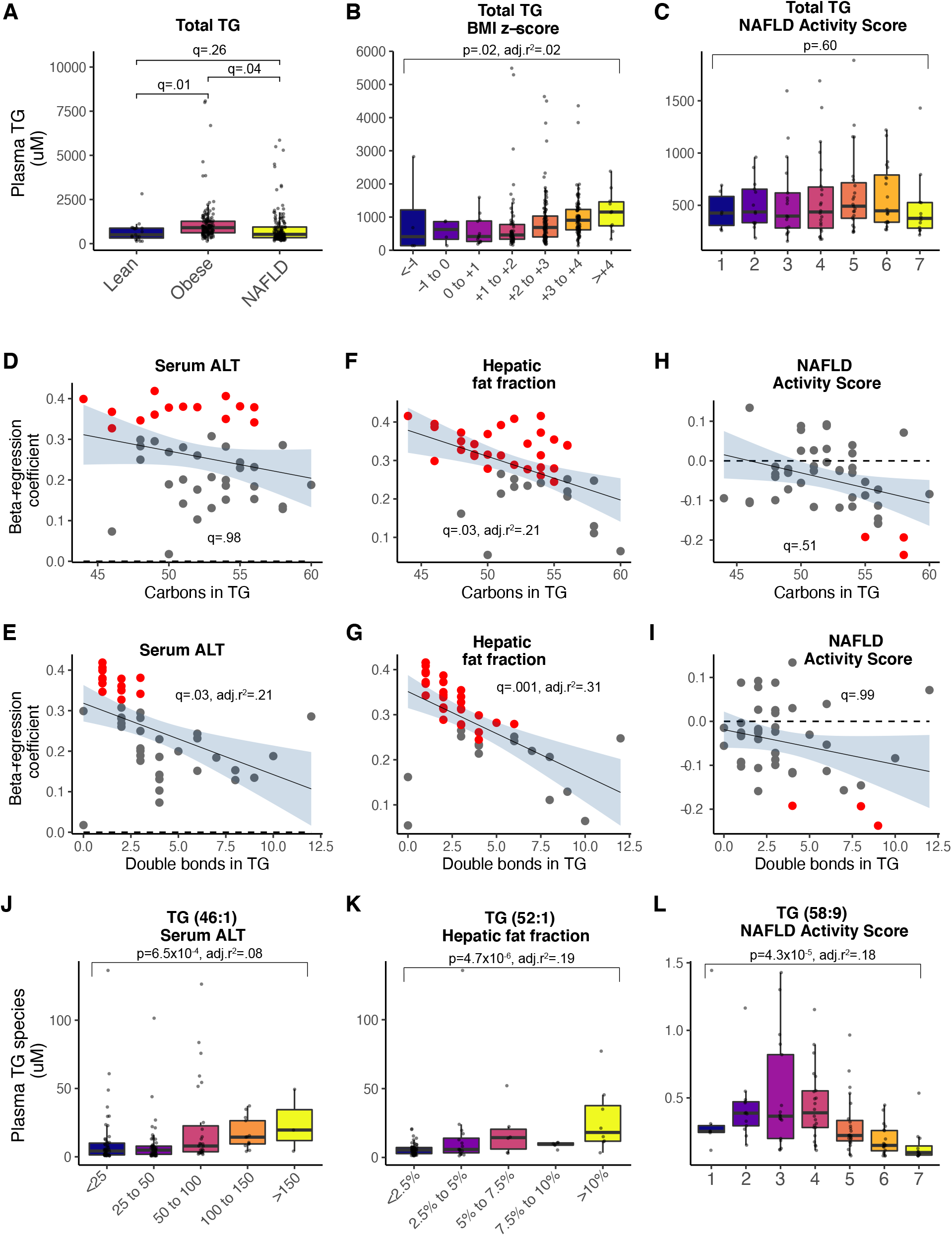
Associations between serum triglyceride species from LC-MS and fatty liver. Total TG was highest in the obese cohort (A) and positively associated with BMI z-score (B), but not associated with NAFLD Activity Score. Results from meta-regression are shown in D-I, where each dot represents a different lipid species and those in red are lipids significantly associated with the trait on linear regression. In the obese cohort (n=146), TG chain length (D) and saturation (E) were negatively associated with serum ALT. In obese children who underwent MRS (n=95), TG chain length (F) and saturation (G) were also negatively associated with hepatic fat fraction. Very long-chain polyunsaturated TG were negatively associated with NAFLD Activity Score in children biopsied for NAFLD (H, I, & L). Whereas, positive associations were observed for short, mono-unsaturated TG with serum ALT (J) and hepatic fat fraction (K) in obese children. Panels B, C, J-L show results from linear regression used log-transformed lipid concentrations adjusted for age and sex, and give p-values where significance is p<3.3×10^−3^. Panels D-I show results from meta-regression, trend line is accompanied by a grey shaded area to illustrate the 95% confidence interval and q-values (derived using the Benjamini-Hochberg method) where significance is q<.05.

Though trends on meta-regression were consistent in the both obesity cohort and biopsied NAFLD cases, the associations of individual lipids were affected by the characteristics of the different cohorts. For example, medium-length, mono-saturated TG (e.g. TG(46:1) and TG(52:1)) were positively associated with serum ALT (Figure 3J) and hepatic fat fraction (Figure 3K) in the obesity cohort. Whilst very-long chain, polyunsaturated TG (e.g. TG(58:9), Figure 3L) were negatively associated with NAS in the biopsied NAFLD cases.

Consistent results across biopsied NAFLD cases and the obesity cohort were also observed for phosphatidylcholines (PC). Total PC was lower in the obesity cohort than lean controls and lowest in biopsied NAFLD cases (Figure 4A). Serum ALT and hepatic fat fraction (in obese cohort), and NAS (in NAFLD cohort) were negatively associated with PC desaturation and chain length (Figure 4). Again, the associations of individual lipids were affected by the characteristics of the different cohorts. For example, medium-length, mono-unsaturated PC (e.g. PC(36:1)) were positively associated with serum ALT (Figure 4J) and hepatic fat fraction (Figure 4K) in the obesity cohort. Whereas, long chain, polyunsaturated PC (e.g. PC(38:5), Figure 4L) was negatively associated with NAS in the biopsied NAFLD cases.

**Figure 4.**
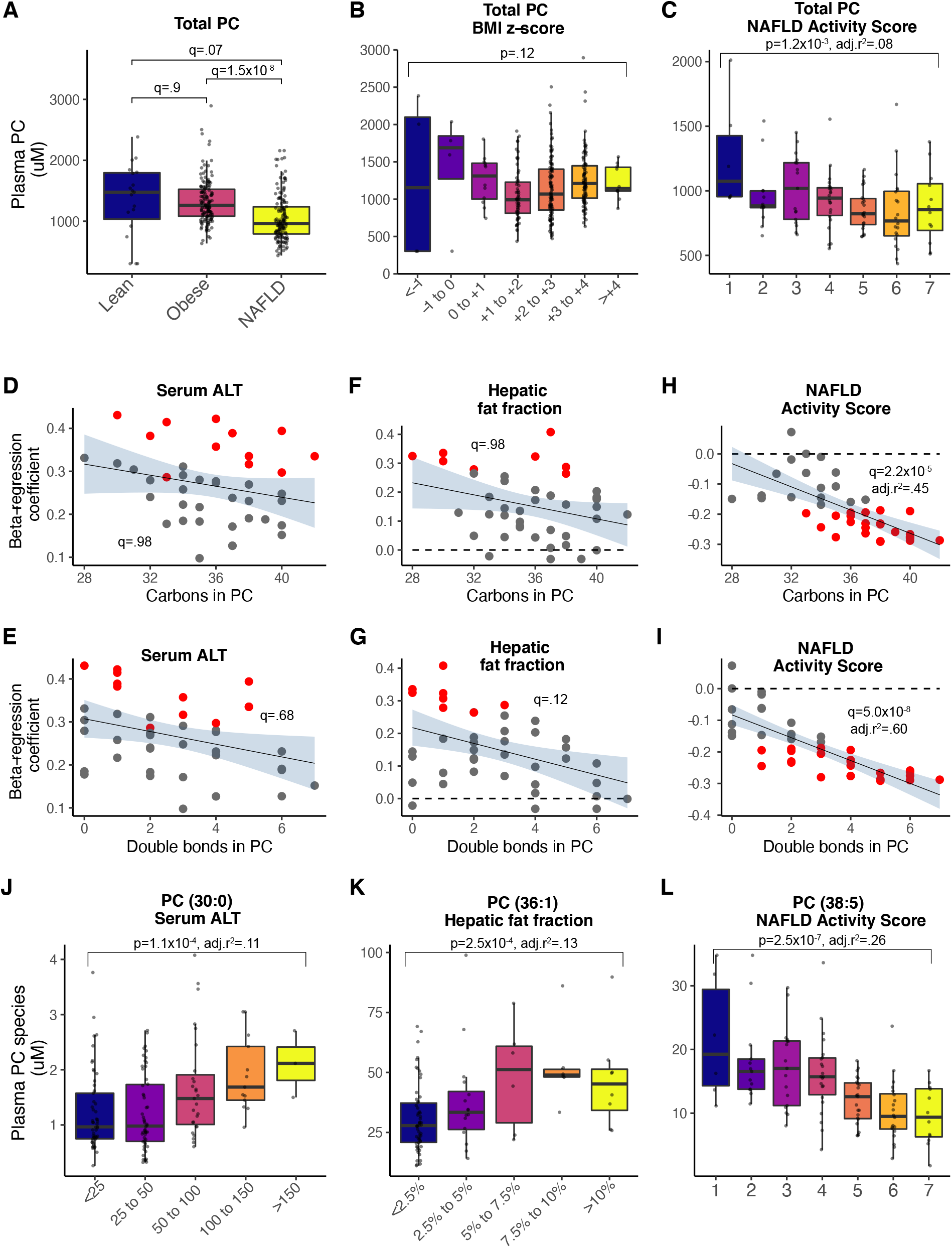
Associations between serum phosphatidylcholine (PC) species from LC-MS and fatty liver. Total PC was lowest in the NAFLD cases (A) and negatively associated with NAFLD Activity Score (C). Results from meta-regression are shown in D-I, where each dot represents a different lipid species and those in red are lipids significantly associated with the trait on linear regression. In the obese cohort (n=146), there was no significant trend on meta-regression between PC chain length (D) and saturation (E) with serum ALT or hepatic fat fraction (n=95, F & G). Long-chain polyunsaturated PC were negatively associated with NAFLD Activity Score in children biopsied for NAFLD (H, I, & L). Whereas positive associations were observed for short, un/mono-unsaturated PC with serum ALT (J) and hepatic fat fraction (K) in obese children. Panels B, C, J-L show results from linear regression used log-transformed lipid concentrations adjusted for age and sex, and give p-values where significance is p<3.3×10^−3^. Panels D-I show results from meta-regression, trend line is accompanied by a grey shaded area to illustrate the 95% confidence interval and q-values (derived using the Benjamini-Hochberg method) where significance is q<.05.

We also observed a positive association between levels of lysophosphatidylcholines (LPC) and severity of NAFLD. Total LPC was higher in the obese cohort than lean controls and highest in the biopsied NAFLD cases (Figure 5A). Saturated LPC (e.g. LPC(16:0) and LPC(18:0), Figure 5C) were positively associated with NAFLD activity score.

**Figure 5.**
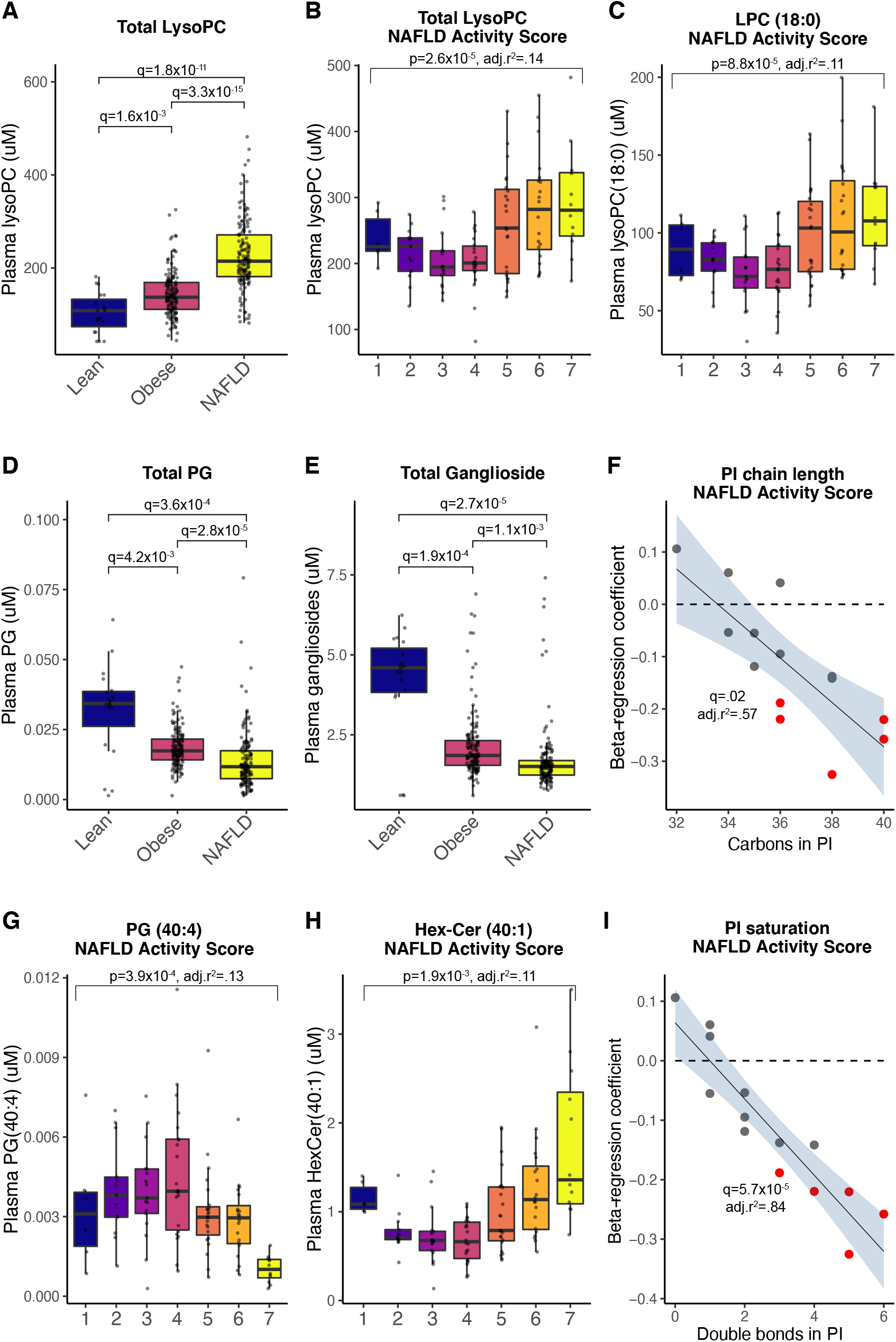
Additional significant associations between serum lipid species from LC-MS and fatty liver in children. Total lyso-phosphatidylcholines (LPC) were highest in the NAFLD cases (A) and positively associated with NAFLD Activity Score (B), particularly for the saturated LPC (e.g. LPC(18:0), C). Whereas total phosphatidylglycerols (PG, D) and total gangliosides (E) were lower in obese children than controls and lowest in the NAFLD cases. Results from meta-regression are shown in F & I, where each dot represents a different lipid species and those in red are lipids significantly associated with the trait on linear regression. Phosphatidylinositol (PI) chain length (F) and saturation (I) were negatively associated with NAFLD Activity Score. Some lipids, such as PG(40:0) (G) and hexosylceramide (40:1) (H) demonstrated non-linear associations with NAFLD activity score. Panels A, D, & E show results from comparison of means (using t-tests) for total lipid abundance for each species. Panels B, C, G, & H show results from linear regression used log-transformed lipid concentrations adjusted for age and sex, and give p-values where significance is p<3.3×10^−3^. Panels F & I show results from meta-regression, trend line is accompanied by a grey shaded area to illustrate the 95% confidence interval and q-values (derived using the Benjamini-Hochberg method) where significance is q<.05.

Several other lipid classes demonstrated significant differences between groups, including phosphatidylglycerols (PG) and total gangliosides, which were lower in the obesity cohort and lowest in biopsied NAFLD cases (Figure 5D). Similar to the results observed for saturation and chain length for PC, we found a negative association for phosphatidylinositols (PI) with NAS (Figure 5F & 4I). We also noted that there were non-linear associations between some lipids species with NAS, for example PG(40:4) (Figure 5G-H).

In total, we identified 72 individual lipids associated with activity (portal inflammation or NAS) or fibrosis stage of pediatric NAFLD (Table S5). We next assessed whether these lipids were associated with hepatic steatosis (and metabolic traits) in a cohort of adults[19]. This identified 9 lipids with significant, consistent effects in this independent study (Table 2). Many of these lipids perturbations were also associated with markers of the metabolic syndrome (e.g. higher BMI, body fat, or HOMA-IR) in this adult cohort (Table S6).

**Table 2.** Summary table of top lipids associated with histological severity of NAFLD with children. Lipids were included if: significantly associated with NAFLD Activity Score in children on linear regression adjusted for age and sex, and showed a directionally consistent association with hepatic steatosis on ultrasound in an independent cohort of adults from a published study (Mann et al., 2020). Note that betas are not directly comparable between the current study and Mann et al. 2020 due to differences in analyses. Several of these lipids were also recently reported to be significantly associated with disease outcomes in adults: (+) indicates a positive / (-) negative association between serum lipid levels and trait from Pietzner et al., 2021. Five lipids have previously been reported to have genome-wide significant loci and these variants are also independently associated with cardiometabolic traits on phenome-wide association studies. ALP, alkaline phosphatase; CVD, cardiovascular disease; LysoPC, lyso-phosphatidylcholine; PC, phosphatidylcholine; TG, triglyceride.

| Lipid | Association with<br>NAS in children<br>(current study) |  | Association with<br>steatosis in adults (from<br>Mann et al. 2020) |  | Associated disease outcomes in<br>adults (from Pietzner et al.<br>2021) | GWAS-significant<br>variants in or near<br>genes | Cardiometabolic traits identified<br>from PheWAS |
| --- | --- | --- | --- | --- | --- | --- | --- |
|  | Beta | p-value | Beta | p-value |  |  |  |
| LysoPC 16:0 | 0.20 | 1.1E-03 | 0.06 | 1.7E-05 | breast_ca (+), glaucoma_hosp<br>(+), nmskin_ca (+) | LIPC, MAF,<br>MFSD2A | IHD, Death from PVD/mesenteric<br>ischaemia/SAH, Metabolic<br>syndrome |
| PC 36:4 | -0.23 | 1.5E-04 | -0.12 | 8.8E-03 | colon_ca (-), dead (-),<br>endmtrl_ca (-), inc_cardfail (-),<br>inc_diabetes (-), inc_resp_asth (-<br>) , inc_resp_copd (-),<br>inc_venous_t (-) | FADS1-2-3,<br>FADS2 | Death from CVD, Serum ALP,<br>Arterial thrombosis, HbA1c,<br>Fasting glucose, Metabolic<br>syndrome, Colorectal cancer, Limb<br>fat |
| PC 37:4 | -0.19 | 9.4E-04 | -0.21 | 1.2E-06 |  | FADS2 | Death from CVD, Serum ALP,<br>Arterial thrombosis, HbA1c,<br>Fasting glucose, Metabolic<br>syndrome, Colorectal cancer, Limb<br>fat |
| PC 38:3 | -0.19 | 1.0E-03 | -0.27 | 1.2E-09 |  | FADS1-2-3 | HbA1c, Fasting glucose,<br>Metabolic syndrome, Colorectal<br>cancer, Limb fat |
| PC 38:5 | -0.29 | 2.5E-07 | -0.22 | 1.4E-06 | dead (-), inc_resp_asth (-) | FADS2 | Death from CVD, Serum ALP, Arterial thrombosis, HbA1c, Fasting glucose, Metabolic syndrome, Colorectal cancer, Limb fat |
| PC 38:6 | -0.29 | 4.5E-07 | -0.06 | 3.3E-06 | dead (-), dead (-), inc_cardfail (-), inc_pad_all (-), inc_resp_asth (-), inc_resp_asth (-), inc_resp_copd (-), inc_resp_copd (-) |  |  |
| PC 40:2 | -0.19 | 2.0E-03 | -0.31 | 6.2E-12 |  |  |  |
| PC 40:7 | -0.29 | 5.8E-07 | -0.23 | 3.8E-07 | breast_ca (-), dead (-), inc_diabetes (-), inc_resp_asth (-), inc_resp_copd (-), lung_ca (-) |  |  |
| TG 58:9 | -0.24 | 4.3E-05 | -0.09 | 4.2E-02 |  |  |  |

In order to understand the clinical significance of these lipids we used data from a metabolite-wide association study of non-communicable diseases in adults[20]. We found significant, directionally consistent associations between lipid levels and cardiometabolic disease (Table S6), which are the two top causes of mortality in adults with NAFLD[2]. For example, levels of PC(36:4) were negatively associated with NAS in children (beta -.23, p=.0001), hepatic steatosis in adult (beta -.12, q=.009), cardiac failure (beta -.06, q=.04), and death (beta -.05, q=.0005).

Next, to understand which genes may influence the perturbations we have observed, we identified published GWAS-significant loci[21] associated with these lipids. We found variants in or near 12 genes at GWAS-significance with these lipids, with the most variants in or near FADS1-2-3 (Table S7). For example, levels of PC(36:4) were associated with rs174564A>G near FADS1-2-3 (beta -.62, p=8.1×10^−72^).

Finally, to provide further clinical context, we identified cardiometabolic traits associated with these variants (Table S8). Variants in or near *FADS1-2-3, LIPC, MAF*, and *MFSD2A* were associated with (death from) cardiometabolic disease, higher fasting glucose, and body fat (Table 2). For example, lower plasma PC(36:4) was associated with: higher NAS in children; hepatic steatosis in adults (from Mann *et al*.); all-cause mortality, diabetes, and cardiac failure in adults (from *Pietzner et al*.); and, multiple variants in *FADS1-2-3*, which are also independently linked to death from cardiovascular disease and insulin resistance (Table 2).

## Discussion

There is a well-established association between paediatric NAFLD, insulin resistance, and obesity in childhood but the long-term metabolic outcomes of this condition have not yet been fully described. Moreover, due to differences in phenotype, it has been unclear whether children share the same perturbations of lipid metabolism as adults with NAFLD. Therefore, we examined the serum lipid profile of obese children and those with NAFLD to improve our understanding of their lipid metabolism, then compared findings with the results of studies in adults. We found that several lipid species associated with insulin resistance and NAFLD severity in children were also linked to hepatic steatosis and cardiometabolic disease outcomes in adults.

Our analysis identified perturbation of multiple lipid groups, including PC and LPC. The majority of other lipidomic studies in NAFLD have also identified substantial associations between PC species and NASH. Whilst much of this work to date has been in adults[12,13,31,32], the similarities between findings suggest similar alterations in lipid metabolism between adult and paediatric NAFLD.

Phosphatidylcholines are membrane-forming lipids and therefore in circulation their abundance is influenced by the concentration of lipoprotein particles, which outer shell consists mainly of PCs. Hartley *et al*. observed lower concentrations of large- and medium-sized HDL in children with NAFLD, which could account for lower PC[18]. We also found higher levels of several saturated LPC to associated with NAFLD severity, similar to the findings by Puri *et al*. (in the livers of adults with NASH)[11]. In addition, LPC(18:1) was one of the top species identified in a separate study that differentiated obese controls from children with NAFLD[17]. The LPC identified in the present study (LPC(16:0, 18:0)) are consistent with the activity of phospholipase A_2_ (PLA_2_) on PCs[33], therefore increased PLA_2_ activity might account for these observations. There are several potential sources of increased PLA_2_ activity, including lipoprotein-associated PLA_2_ (Lp-PLA_2_) and secreted forms of PLA_2_ [34]. PLA_2_ is of particular interest in NAFLD (and cardio-metabolic disease) as its activity is thought to correlate with pro-inflammatory mediators, and presence of oxidized-LDL[35,36]. Whether (paediatric) NAFLD is an independent risk factor for atherosclerosis is a complex question[37], however cardiovascular disease is the top cause of mortality in NAFLD[2]. We found some lipids that are not routinely measured in plasma or serum (like gangliosides and phosphoglycerols (PGs). Work on cells and tissue samples suggests that these are necessary for normal hepatocyte structure[38] and changes in the circulation reflect what is happening at a tissue level[32]. We found that the other lipids are consistently associated with cardiovascular disease across multiple cohorts and via genetic variants influencing these lipids, though the direction of causality remains unclear.

There is a strong body of work implicating hepatic *de novo* lipogenesis (DNL) is upregulated in NAFLD[39] and is both a cause and a consequence of insulin resistance[40,41]. We observed a strong correlation between liver fat and HOMA-IR in both groups. Lipidomic research has suggested that specific lipids can serve as indirect evidence for changes in DNL[42]. Hepatic DNL is classically associated with a higher abundance of short-chain, saturated TG and lower, long-chain, unsaturated TG[43], which correlates closely with the effects of (hepatic) insulin resistance[44]. This had been observed in a pilot study on obese teenagers[45] and we replicated these findings across both our obesity cohort and paediatric biopsied NAFLD cases. Overall, we consider these results to be reflective of hepatic insulin resistance but an alternative methodology, for instance using isotopically labeled substrates, would be needed to formally investigate DNL in paediatric NASH.

A wide range of associations have been identified in previous metabolite (or lipid) profiling studies in children with NAFLD. Several have found higher levels of (branched-chain) amino acids[17,18,46–50]. There is strong evidence that levels of circulating amino acids, particularly branched chain amino acids, are highly correlated to (and even causal of) insulin resistance[51,52]. Other studies have focused on gastro-intestinal tract-derived metabolites[53], which also appear to show utility in differentiating controls from NAFLD, though are less effective in separating NAFL and NASH[54]. We did not attempt to use our data to derive a prediction algorithm due to the lack of a second, independent cohort of children biopsied for NAFLD.

These findings underscore the importance of interventions that reverse insulin resistance and dyslipidaemia in this group of children. Long-term exposure of their vasculature to this lipid profile may lead to premature cardiovascular disease. Therefore, clinical measures (i.e. weight loss) in children with NAFLD that normalize their serum lipid profile are likely to be of benefit if sustained long-term.

The main strength of this study was the inclusion of participants from a spectrum of severity of paediatric metabolic syndrome. This facilitated demonstration of trends in two separate groups of children with insulin resistance. We also were able to use histological severity of NAFLD as our main outcome, which is the gold standard of assessment and comparatively few liver biopsies are performed for fatty liver in children. Lastly, use of multiple publicly available datasets provided supporting clinical context for top lipids and comparison with results from adults, this shows that the international lipidomics community is providing a strong evidence base for new studies to build on.

The principal limitation of this work is the lack of a second, biopsied cohort of children with NAFLD for validation of results. In addition, liver biopsy samples were not available for lipidomics, which would have improved our understanding of lipid metabolism at the level of the hepatocyte. However, studies that have included paired liver and plasma samples have found considerable overlap between significantly associated species[13]. We also identified several changes in the plasma consistent with previous reports from liver samples in adults[11]. Given the strong correlation between steatosis grade and NAS, and that comparatively few children in this cohort had severe fibrosis, these results are most informative of mild-moderate NAFLD driven by liver fat content. It should also be noted that our participants were primarily of non-Finnish European descent and therefore it is unclear to what extent these findings are generalisable to other ethnicities. Whilst we have illustrated several lipids of interest associated with the severity of paediatric NAFLD, further work, both practical and conceptual, would be required to validate these findings and progress this technique towards clinical utility. Lastly, as a cross-sectional study, we are unable to determine causality or define specific mechanisms for alterations of lipids.

## Conclusion

Severity of paediatric NAFLD and insulin resistance are inversely associated with long-chain, polyunsaturated PC and TG, and positively associated with saturated LPC. These lipids are also linked to hepatic steatosis, insulin resistance, and cardiometabolic disease outcomes in adults. Pediatric NAFLD shows similar perturbation of lipid metabolism to the metabolic syndrome in adults, therefore reversal of insulin resistance in these patients is needed to reduce long-term complications.

## Supporting information

Graphical Abstract

## Data Availability

Full summary statistics available in supplement. Code used in analysis available from: https://doi.org/10.5281/zenodo.5507129

## Abbreviations

GWAS: genome-wide association study
LPC: lyso-phosphatidylcholine
NAFLD: non-alcoholic fatty liver disease
NASH: non-alcoholic steatohepatitis
NAS: NAFLD Activity Score
PC: phosphatidylcholine
PheWAS: phenome-wide association study
TG: triglyceride

## Data availability

All data and results from analyses are contained within the manuscript and supplement. Code used in analyses is available from https://doi.org/10.5281/zenodo.4656980.

## Acknowledgements

The authors are grateful to the young people and their families for taking part in the EU-PNAFLD Registry. The other EU-PNAFLD investigators are: Piotr Socha, Wojciech Jańczyk, Ulrich Baumann, Donatella Comparcola, Sanjay Rajwal, Florence Lacaille, Myriam Dabbas, Quentin M. Anstee and the late Valerio Nobili.

## Supplementary data

**Table S1. Summary of baseline histology for children biopsied for NAFLD (n=122)**. Histology was scored according to the NASH CRN system.

**Table S2. Associations between histology and traits for children biopsied for NAFLD (n=122)**. Beta represents the linear regression coefficient between histological outcome and baseline trait. Significance was defined as q<0.05, using Benjamini-Hochberg method for FDR correction from p-values.

**Table S3. Full results of associations between lipids (n=229) and traits**. Beta represents the linear regression coefficient between log-normalized serum lipid concentration and trait. Significance was defined as p<0.5(/sqrt(n)), where n=number of lipids, due to high correlation between lipid species. Traits studies: ALT, serum alanine aminotransferase in the obese cohort (n=146); TE, transient elastography in the obese cohort (n=42); Fib, fibrosis stage on biopsy in the NAFLD cohort (n=122); Steat, histological steatosis severity in the NAFLD biopsied cohort (n=122); Portal, portal inflammation severity in the NAFLD biopsied cohort (n=122); NAFLD_HOMA, HOMA-IR in the NAFLD biopsied cohort (n=122); Ob_HOMA, HOMA-IR in the obese cohort (n=146); NAS, NAFLD Activity Score in the NAFLD biospied cohort (n=122).

**Table S4. Full results of meta-regression analyses between lipids saturation or chain length and traits**. Beta represents the regression coefficient between lipid chain length (‘Carb’) or saturation (‘db’, for number of double bonds) and traits. Significance was defined as q<0.05, using Benjamini-Hochberg method for FDR correction from p-values. Traits studies: Ob_HOMA, HOMA-IR in the obese cohort (n=146); Ob_HFF, hepatic fat fraction on magnetic resonance imaging in the obese cohort (n=95); Ob_TE, transient elastography in the obese cohort (n=42); NAFLD_NAS, NAFLD Activity Score in the NAFLD biospied cohort (n=122); NAFLD_Steat, histological steatosis severity in the NAFLD biopsied cohort (n=122); NAFLD_Fib, fibrosis stage on biopsy in the NAFLD cohort (n=122); NALD_Portal, portal inflammation severity in the NAFLD biopsied cohort (n=122); NAFLD_HOMA, HOMA-IR in the NAFLD biopsied cohort (n=122); Ob_ALT, serum alanine aminotransferase in the obese cohort (n=146).

**Table S5. Serum lipids significantly associated with histological severity of NAFLD in children**. Results from linear regressions log-normalized serum lipid concentration and traits. Significance was defined as p<0.5(/sqrt(n)).

**Table S6. Full results of overlap between lipids significantly associated with histological NAFLD in children from the current study and phenotypes in adult cohorts**. Using data from two published metabolomics studies (Mann et al. Hum Mol Genet (2020), and Pietzner et al. Nat Med (2021)) we filtered associations between lipids and traits with false-discovery rate corrected q-values of <0.1. Overlap was identified as significantly associated with lipids from the current study (Table S5) had a directionally consistent beta regression coefficient in the adult cohort. Traits assessed in Mann et al. Hum Mol Genet (2020): hepatic steatosis on abdominal ultrasound without (‘Steatosis’) and with adjusting for serum markers of insulin resistance (‘Steatosis_IR_adjust’); association between lipids and variants in or near genes known to be associated with severity of NAFLD in adults (MTARC1, GCKR, MBOAT7, PNPLA3, TM6SF2, SERPINA1, PPP1R3B); insulin resistance (IR); body mass index (BMI); BMI-adjusted waist-to-hip ratio (WHRadjBMI); total body fat (BodyFat). Traits assessed in Pietzner et al. Nat Med (2021): breast cancer (breast_ca), cataracts (cataract_hosp), colonic carcinoma (colon_ca), death (dead), endometrial carcinoma (endmtrl_ca), fracture (frac_all_hosp), glaucoma (glaucoma_hosp), abdominal aortic aneurysm (inc_aaa), atrial fibrillation (inc_af), cardiac failure (inc_cardfail), cerebrovascular infarction (inc_cvainfct), diabetes (inc_diabetes), ischaemic heart disease (inc_ihd), liver disease (inc_liver), peripheral artery disease (inc_pad_all), kidney disease (inc_renadis), asthma (inc_resp_asth), chronic obstructive pulmonary disease (inc_resp_copd), venous thrombo-embolism (inc_venous_t), lung cancer (lung_ca), non-melanoma skin cancer (nmskin_ca), oesophageal cancer (oesoph_ca), rectal carcinoma (rectum_ca), and stomach carcinoma (stomach_ca).

**Table S7. Previously published genome-wide significant loci for lipids associated with histological severity of NAFLD in children**. Using data from published lipidomics GWAS we identified GWAS-significant loci (as defined by each contributing study) for the lipids listed in Table S5.

**Table S8. Full results from a phenome-wide association study (PheWAS) for GWAS-significant variants from Table S7**. Cardiometabolic traits were extracted from Phenoscanner_v2 and Tabassum et al. (2019) for each variant in Table S7, which corresponded to a lipid from Table S5.

**Figure S1. Characteristics of obesity cohort and cases biopsied for NAFLD**. Obesity cohort (n=146) blue) was collected from any child who was overweight or obese referred for clinical assessment. The NAFLD cases (n=122, pink) were children with suspected severe paediatric NAFLD who underwent liver biopsy. There is no data shown from the lean group. The two overlying histograms illustrate the distribution of traits for body mass index (BMI) z-score (A), homeostatic model of insulin resistance (HOMA-IR, B), serum triglycerides (C), serum total cholesterol (D), and serum alanine aminotransferase (ALT, E). A subset (n=95) children from the obesity cohort underwent magnetic resonance spectroscopy (MRS) for estimation of liver fat (F). q-values represent comparison of means using t-test derived using the Benjamini-Hochberg method where significance is q<.05.

**Figure S2. Associations between baseline characteristics and severity of peri-portal inflammation in children biopsied for NAFLD (n=122)**. None of the baseline characteristics demonstrated an association with severity of portal inflammation. Associations were tested using linear regression. q-values were derived using the Benjamini-Hochberg method where significance is q<.05.

**Figure S3. Summary of serum lipidomics by LC-MS performed on 287 children**. After normalization, there was no difference between lean (n=19), obese (n=146), and NAFLD (n=122) groups on principal component analysis (A). Lipids from each class were correlated using hierarchical clustering (B) but there was no clear distinction between obese and NAFLD groups. (C) Over 50 lipid species were significantly associated with individual traits within either the obese or NAFLD groups using linear regression adjusted for age and sex. The cell color on represents the beta regression coefficient for each analysis and stars illustrate p-values, where significance is p<3.3×10^−3^. On meta-regression (D), lipid saturation and carbon chain length were significantly associated with several traits in both the obese and NAFLD groups. The cell color in (D) represents the meta-regression beta coefficient and stars illustrate q-values (derived using the Benjamini-Hochberg method) where significance is q<.05.

