## Supplementary figures and images for "Plasma lipidomics identifies a signature of NAFLD in children that couples with cardiometabolic outcomes in adults"

### FigS1

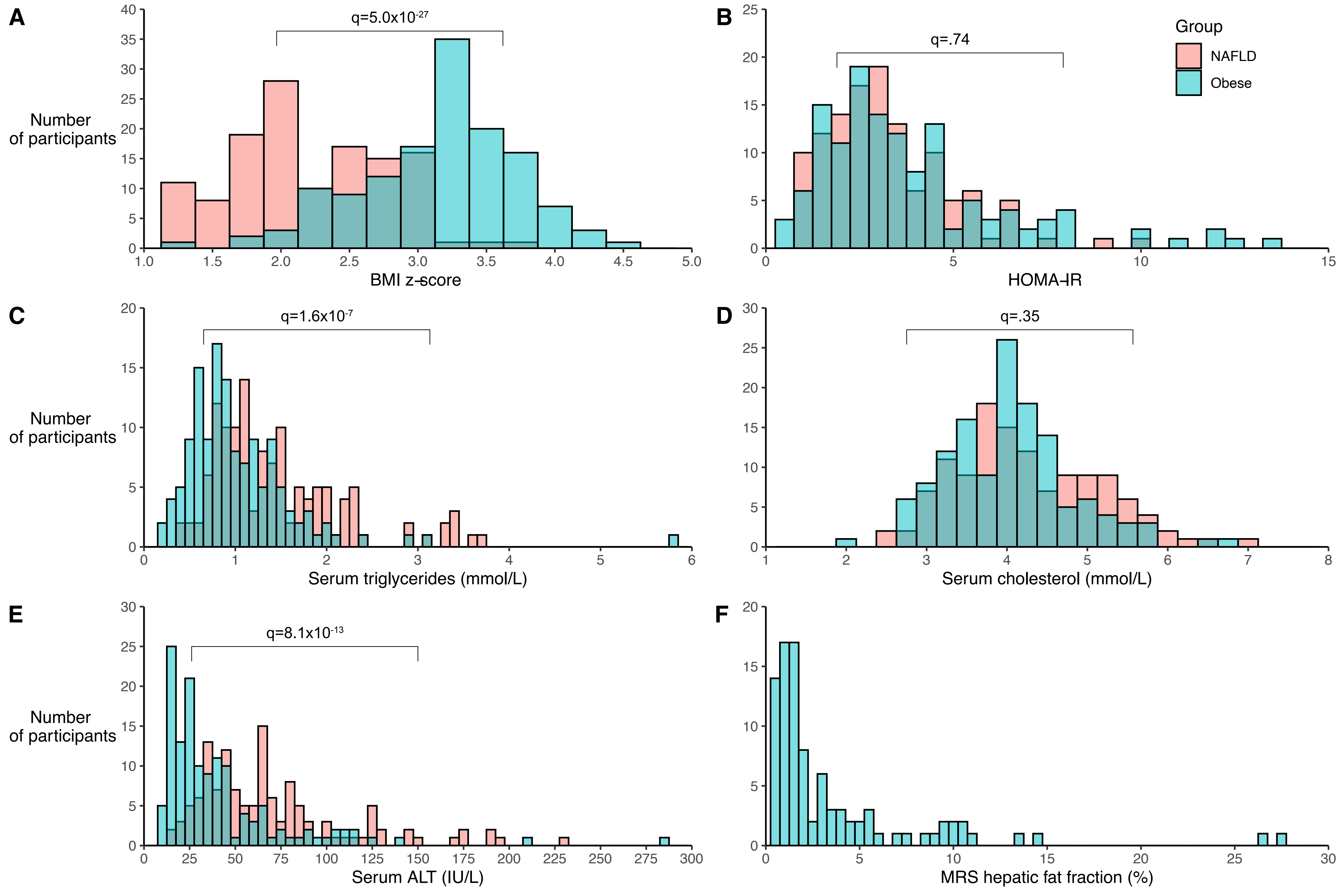

### FigS2

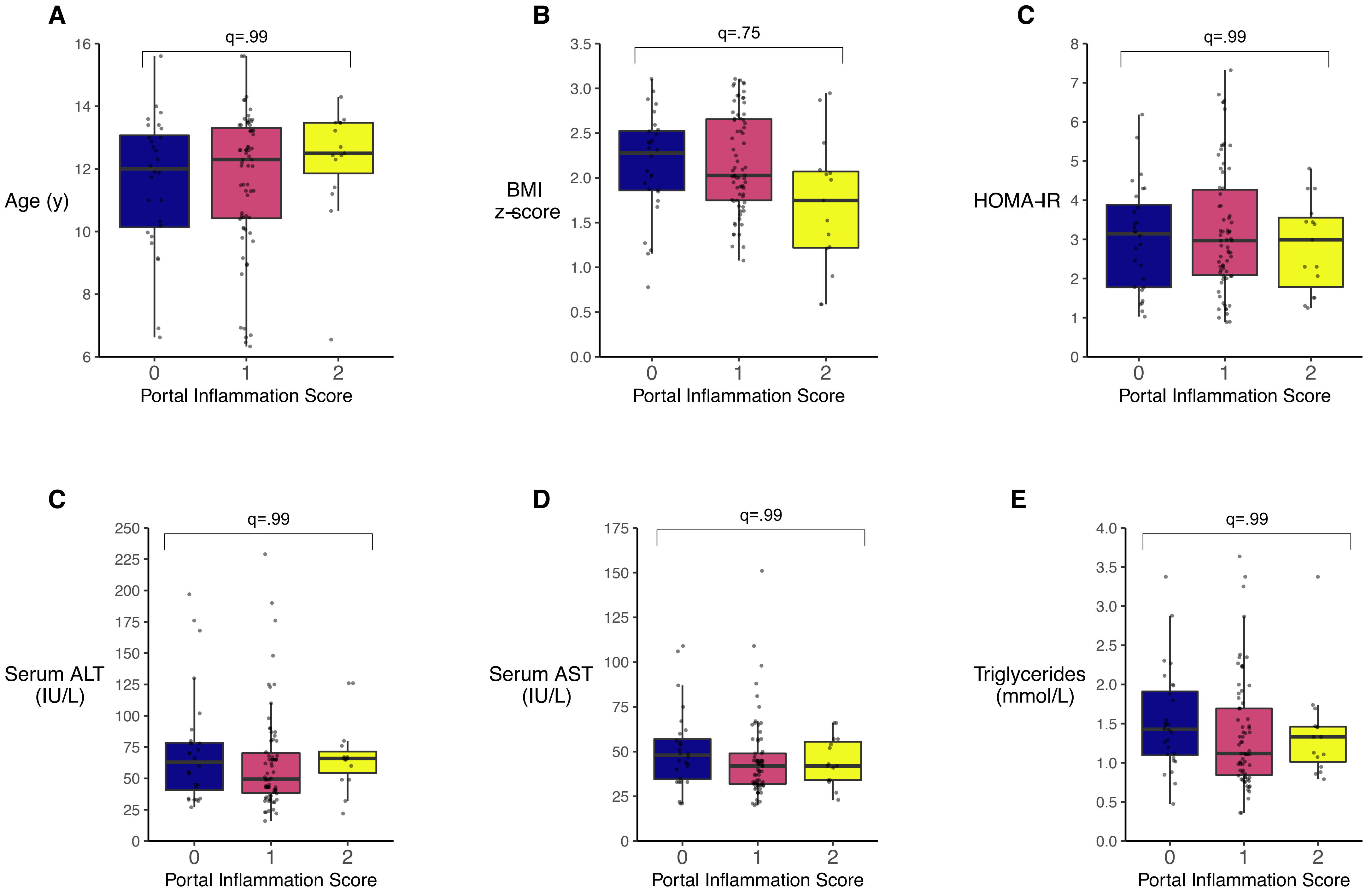

### FigS3

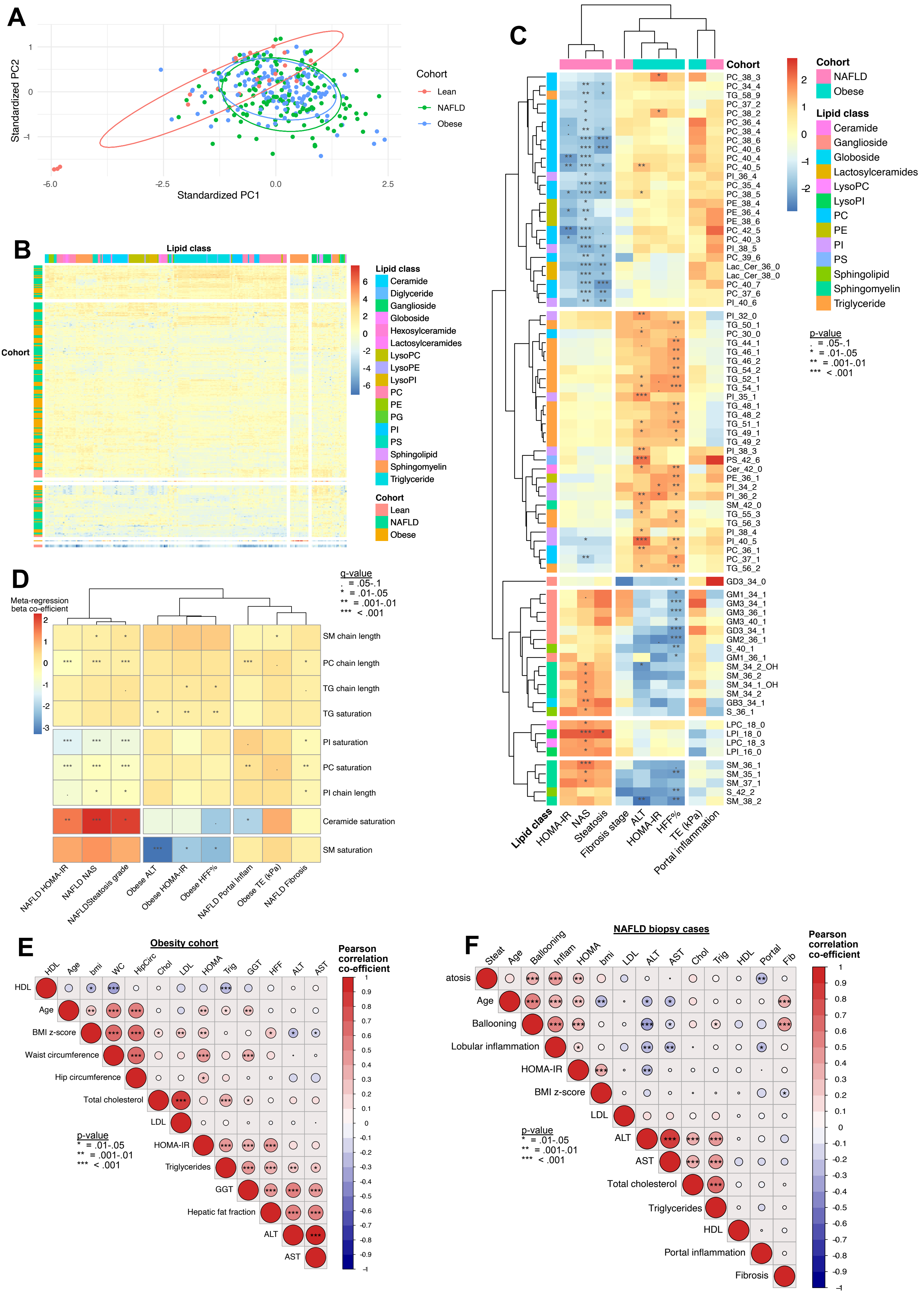

### Graphical Abstract

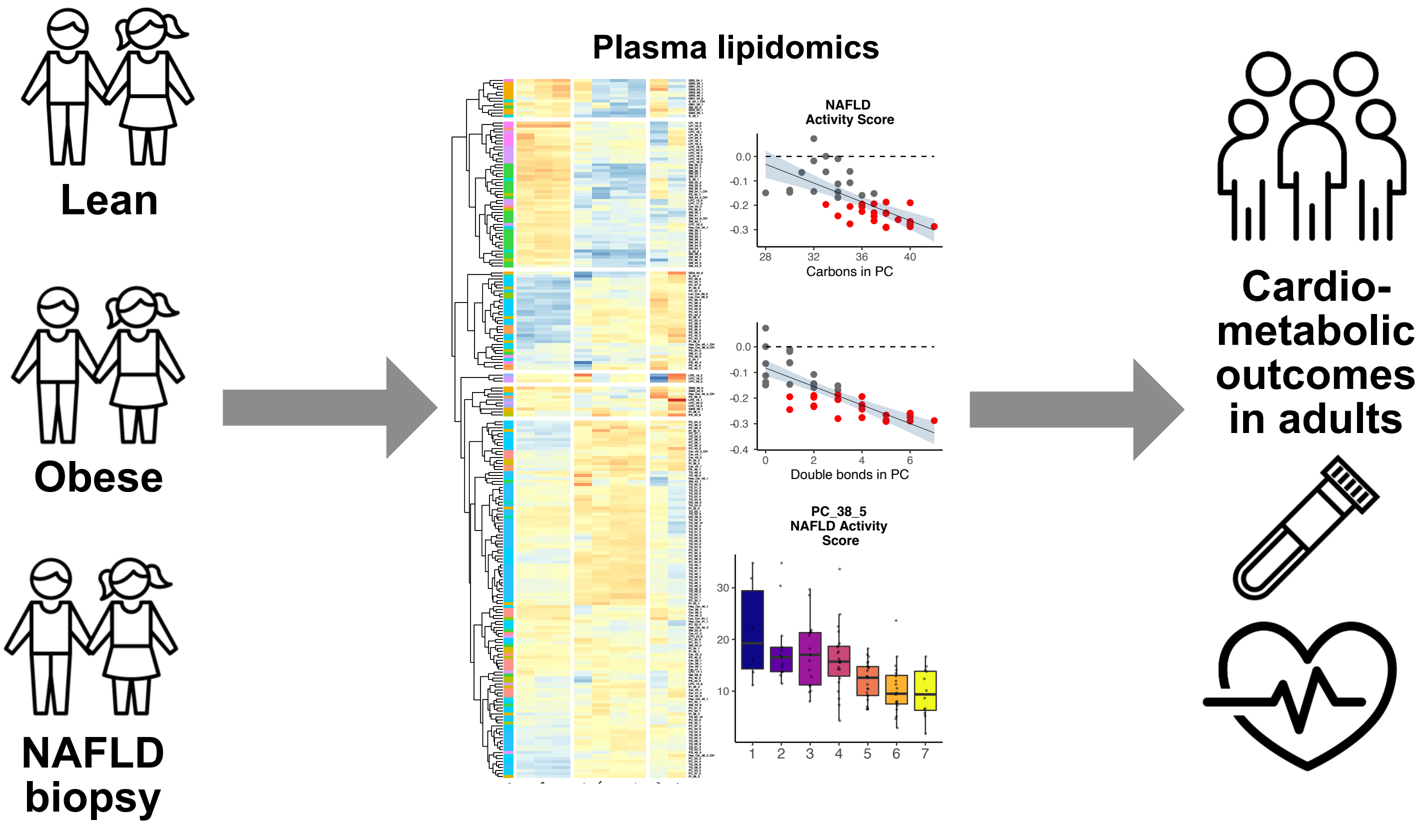
